# Estimating the incubation period of monkeypox virus during the 2022 multi-national outbreak

**DOI:** 10.1101/2022.06.22.22276713

**Authors:** Kelly Charniga, Nina B. Masters, Rachel B. Slayton, Lucas Gosdin, Faisal S. Minhaj, David Philpott, Dallas Smith, Shannon Gearhart, Francisco Alvarado-Ramy, Clive Brown, Michelle A. Waltenburg, Christine M. Hughes, Yoshinori Nakazawa

## Abstract

Monkeypox is a zoonotic disease endemic in Central and West Africa. In May 2022, an outbreak of monkeypox characterized by human-to-human transmission was detected in multiple non-endemic countries. We estimated the incubation period for monkeypox using information from 22 probable (N = 1) and confirmed (N = 21) monkeypox cases in patients reported in the United States through June 6, 2022. We pooled U.S. patient data with the data from 18 confirmed cases in patients reported from the Netherlands through May 31, 2022. The mean incubation period from exposure to first symptom onset was 7.6 days (95% credible interval: 6.2 – 9.7), and the 95th percentile was 17.1 days (95% CrI: 12.7–24.3). These findings align with current CDC recommendations for monitoring close contacts of people with monkeypox for 21 days after their last exposure.

## Background

Monkeypox is a zoonotic disease endemic in Central and West Africa. Between January 1, 2022 and June 10, 2022, 1536 suspected monkeypox cases were reported in the WHO African Region, with 72 deaths^1^. From May 13, 2022 to June 10, 2022, monkeypox cases were reported in 28 non-endemic countries across four World Health Organization (WHO) Regions^1^. The United Kingdom has reported the largest number of monkeypox cases, with 574 cases reported by June 16, 2022^2^.

Human-to-human transmission of monkeypox virus (MPXV) occurs through close contact with infectious material from skin lesions, respiratory secretions during prolonged face-to-face contact, and fomites such as linens and bedding^3^. The incubation period for MPXV has historically been reported as 4 – 14 days^4^ but could be as long as 17 days^5^. Monkeypox symptoms classically begin with a prodrome (e.g., fever, headache, muscle aches, fatigue, lymphadenopathy). Within one to three days after fever onset, the patient develops a rash, which progresses from macules to papules to vesicles to pustules, and then scabs over^6^. Patients are considered infectious until all lesions have resolved, the scabs have fallen off, and a fresh layer of intact skin has formed.

There is evidence that the incubation period may depend on the route of transmission^7^. For example, data from the 2003 monkeypox outbreak in the United States demonstrated that people who were exposed to MPXV by non-invasive routes, such as petting an infected animal, experienced slower illness progression and longer incubation period than those with complex exposures, such as a bite or scratch from an infected animal that broke the skin. Compared to the classical disease presentation, patients in the 2022 multi-national monkeypox outbreak have had variable clinical presentation, often presenting with a smaller number of lesions, lesions originating in the genital or perineal/perianal area, and lesions appearing before the onset of other symptoms^1,8^. It is important to estimate the incubation period of MPXV for the current outbreak to better understand and tailor public health messaging and interventions.

We estimate the incubation period of MPXV using data from monkeypox patients reported through June 6, 2022, where the exposure period and symptom onset date were known.

## Methods

### Population

From May 17, 2022 to June 6, 2022, 30 probable and confirmed cases of monkeypox were reported in the United States. Data from eight patients were excluded from this analysis due to unknown or uncertain information on window of exposure or symptom onset. Data from 22 probable (N=1)^a^ and confirmed (N=21)^b^ monkeypox patients were included in this analysis.

### Incubation period estimation

We estimated the incubation period of MPXV from exposure to first symptom onset. As initial symptoms for some patients were non-specific (e.g., fever, diarrhea, headache, sore throat) and may not have necessarily been related to their MPXV infection, we also estimated the incubation period using the date of rash onset.

During case investigations, we obtained the window of exposure to MPXV and both time of first symptom onset and rash onset (Table 1). First symptom onset dates ranged from April 29 to May 27, 2022. For 21 monkeypox patients with available information on rash onset, dates of rash onset ranged from May 1 to June 3, 2022.

**Table 1.** Probable and confirmed monkeypox cases in the United States included in the incubation period analysis (N = 22). Incubation period was defined as exposure until first symptom onset.

| Case number | Exposure window (days) | Days to first symptom onset<br>(min – max) |
| --- | --- | --- |
| 1 | 1 | 4 – 6 |
| 2 | 8 | 0 – 8 |
| 3 | 21 | 1 – 23 |
| 4 | 2 | 2 – 5 |
| 5 | 2 | 3 – 6 |
| 6 | 13 | 0 – 14 |
| 7 | 1 | 9 – 11 |
| 8 | 23 | 0 – 24 |
| 9 | 1 | 10 – 12 |
| 10 | 4 | 1 – 6 |
| 11 | 1 | 6 – 8 |
| 12 | 18 | 0 – 18 |
| 13 | 8 | 7 – 16 |
| 14 | 2 | 0 – 2 |
| 15 | 8 | 8 – 17 |
| 16 | 27 | 1 – 29 |
| 17 | 7 | 2 – 10 |
| 18 | 4 | 1 – 6 |
| 19 | 23 | 0 – 23 |
| 20 | 1 | 1 – 3 |
| 21 | 4 | 3 – 8 |
| 22 | 12 | 0 – 12 |

**Table 2.** Confirmed monkeypox cases in the Netherlands included in the incubation period analysis (N = 18). Incubation period was defined as exposure until symptom onset.

| Case ID | Exposure window (days) | Days to symptom onset (min-max) |
| --- | --- | --- |
| 23 | 1 | 5 – 7 |
| 24 | 1 | 8 – 10 |
| 25 | 1 | 7 – 9 |
| 26 | 1 | 7 – 9 |
| 27 | 1 | 5 – 7 |
| 28 | 1 | 5 – 7 |
| 29 | 1 | 13 – 15 |
| 30 | 1 | 18 – 20 |
| 31 | 7 | 0 – 7 |
| 32 | 11 | 4 – 16 |
| 33 | 5 | 14 – 20 |
| 34 | 1 | 6 – 8 |
| 35 | 1 | 4 – 6 |
| 36 | 1 | 5 – 7 |
| 37 | 11 | 5 – 17 |
| 38 | 1 | 11 – 13 |
| 39 | 1 | 7 – 9 |
| 40 | 8 | 1 – 10 |

**Table 3.** Probable and confirmed monkeypox cases in the United States included in the incubation period analysis (N = 21*). Incubation period was defined as exposure until rash onset.

| Case number | Exposure window (days) | Days to rash onset (min – max) |
| --- | --- | --- |
| 1 | 1 | 4 – 6 |
| 2 | 10 | 0 – 11 |
| 3 | 21 | 6 – 28 |
| 4 | 2 | 2 – 5 |
| 5 | 2 | 3 – 6 |
| 6 | 13 | 1 – 15 |
| 7 | 1 | 9 – 11 |
| 8 | 23 | 0 – 24 |
| 9 | 1 | 10 – 12 |
| 10 | 4 | 7 – 12 |
| 11 | 1 | 6 – 8 |
| 12 | 18 | 0 – 18 |
| 13 | 8 | 7 – 16 |
| 14 | 7 | 1 – 9 |
| 15 | 8 | 9 – 18 |
| 16 | 27 | 8 – 36 |
| 17 | 7 | 2 – 10 |
| 18 | 4 | 2 – 7 |
| 20 | 1 | 11 – 13 |
| 21 | 4 | 13 – 18 |
| 22 | 14 | 2 – 17 |

\*Date of rash onset was missing for case 19.

Whenever possible, we used the exact timing of self-reported exposure. If unavailable, we used information such as length of stay in a country reporting cases to bound the window of exposure. We bounded the time of MPXV infection by the earliest and latest potential windows of exposure, following Lessler et al.^9^.

For the analysis of the incubation period using first symptom onset, we included data from 18 additional confirmed monkeypox patients reported in the Netherlands^10^ for a total sample size of 40 monkeypox patients. Rash onset for these cases was not made publicly available, and thus those data were not included in the estimation of incubation period using rash onset.

### Statistical Methods

We constructed a doubly censored dataset for the incubation period and fitted the distribution using the methods described in Lessler et al. and Reich et al.^9,11^. We assumed the incubation period of MPXV followed a log-normal distribution and used the Metropolis-Hastings Markov chain Monte Carlo algorithm for calibration. This method has been used to estimate the incubation periods of other viruses including Zika virus^9^ and Mayaro virus^12^.

All analyses were conducted in R version 4.1.1 (Vienna, Austria), and we used the R package rjags version 4-13 (https://CRAN.R-project.org/package=rjags).

## Results

### Population

All 22 U.S. monkeypox patients included in this analysis were assigned male sex at birth. They had a median age of 37 years (range 28 – 61). Commonly reported symptoms during the course of illness included lesions in the anal and genital areas, swollen lymph nodes, rectal pain, headache, and fatigue. Only 10 out of 21 (48%) monkeypox patients reported fever (fever was unknown for one patient).

Sixteen out of 22 patients (73%) were likely exposed during international travel, one patient was likely exposed during travel to another U.S. state, and five patients had no history of travel and were linked to other patients locally. All 22 patients reported male to male sexual contact (MMSC).

Demographic and exposure information about the patients from the Netherlands study can be found in Miura et al.^10^.

### Incubation Period

For time from exposure to first symptom onset, we estimated a mean incubation period of 7.6 days (95% credible interval (CrI): 6.2–9.7) (median 6.4, 95% CrI: 5.1 – 7.9) and a standard deviation of 4.9 days (95% CrI: 3.3–8.3) (Figure 1). The 95th percentile was 17.1 days (95% CrI: 2.6–9.3) after exposure.

**Figure 1.**
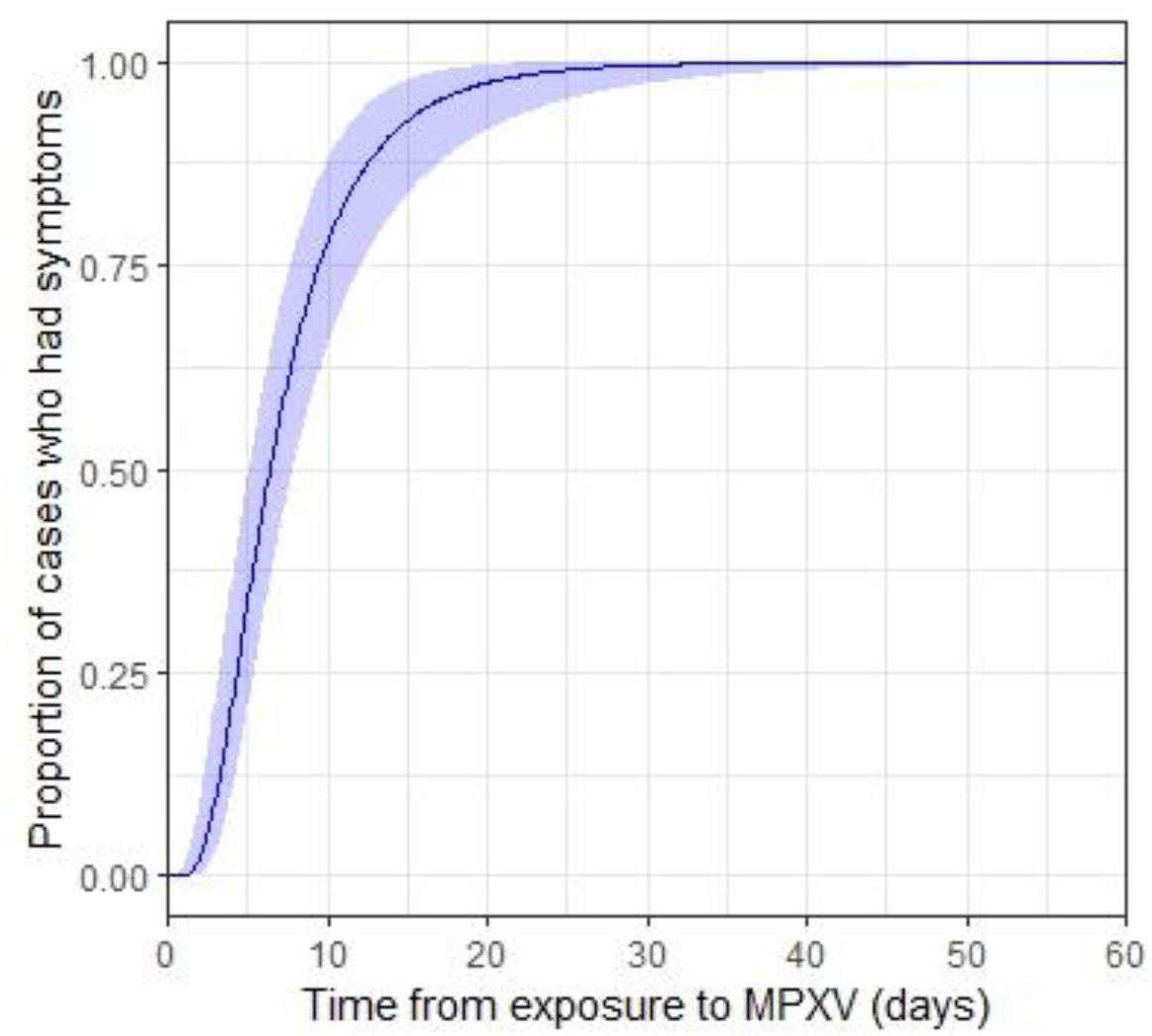
Monkeypox virus incubation period (exposure to **first symptom onset**), United States and the Netherlands, May 2020 – June 2022. Blue shaded area represents 95% credible intervals.

For time from exposure to rash onset, we estimated a mean incubation period of 8.7 days (95% CrI: 6.9–11.7) (median 7.8 days, 95% CrI: 5.9 – 10.0) and a standard deviation of 4.3 days (95% CrI: 2.6–9.3) (Figure 2). The 95th percentile was 17.7 days (95% CrI 12.4–28.1) after exposure.

**Figure 2.**
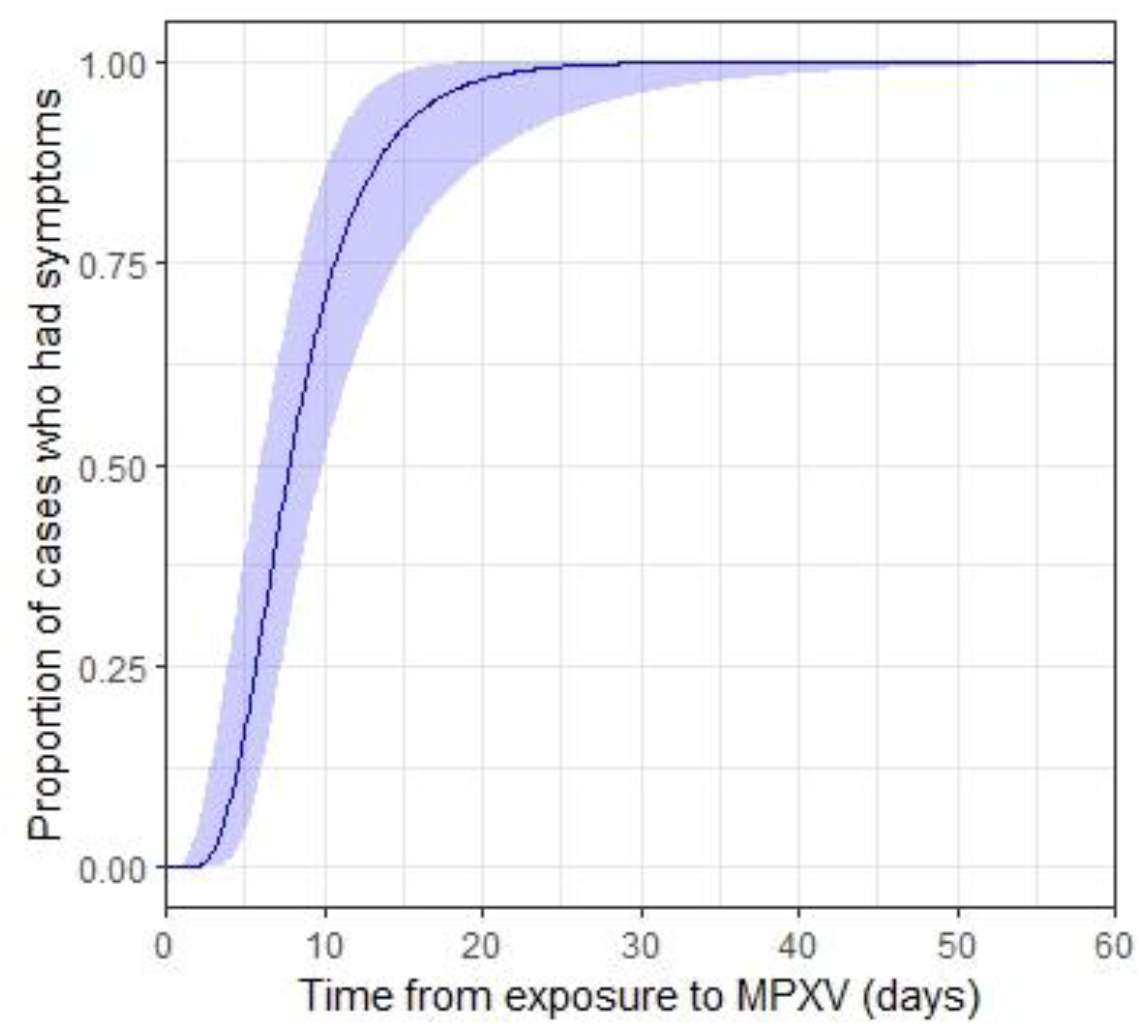
Monkeypox virus incubation period (exposure to **rash onset**), United States, May 2020 – June 2022. Blue shaded area represents 95% credible intervals.

## Discussion

Our estimate of the incubation period from exposure to first symptom onset is shorter than that estimated by Miura et al. (mean 8.5 days, 95% CrI: 6.6 – 10.9)^10^; however, our results are broadly consistent given the uncertainty in the estimates. Our findings that the 95th percentile of the incubation period was 17.1 days supports current guidelines for monitoring people who have been exposed to MPXV for 21 days after last exposure^13^.

The estimated incubation period for this outbreak in the United States and the Netherlands is similar to previous MPXV outbreaks; Nolen et al. reported a mean incubation period of 8 days (range 4–14) for an outbreak of MPXV in the Democratic Republic of Congo in 2013^4^. They defined incubation period as the number of days between contact with a person with symptomatic monkeypox and development of rash. Among 47 cases of monkeypox reported from 1970 – 1979 in Central and West Africa, four instances of secondary transmission may have occurred in families. Onset of rash in secondary cases occurred up to 17 days after the first case^5^. These data support both our estimates and the 21-day monitoring period for contacts of monkeypox cases.

Most monkeypox cases in the United Kingdom have occurred among men (311/314, 99%); of those with available information, 151/152 identified as gay, bisexual, and other men who have sex with men (MSM)^14^. Among the 22 U.S. monkeypox patients included in the incubation period analysis presented in this paper, all were male and reported MMSC. The modes of transmission during sexual contact remain unknown,^8^ but close physical contact with infected persons can spread monkeypox, and thus any person, irrespective of gender or sexual orientation, can acquire and spread monkeypox. The role of bodily fluids in the transmission of monkeypox remains unclear.

As mentioned by Miura et al.,^10^ this report includes early cases reported during this outbreak, and thus people with MPXV infection with a longer incubation period necessarily have a lower probability of inclusion, potentially biasing our estimates of incubation period downward. However, the exposure window has been difficult to ascertain for people who have more recently received a monkeypox diagnosis in the United States.

Interviews with monkeypox patients conducted in the United Kingdom highlight difficulties faced with traditional contact tracing during this outbreak^14^, as most interviewed patients reported sexual contact with new or casual partners for whom contact details were unavailable. This emphasizes the importance of estimating the incubation period to better understand and tailor public health messaging and interventions. Endo et al. highlight that a heavy-tailed (right skewed) partnership distribution over a sexual contact network can explain the rapid growth of transmission observed in the current multi-national monkeypox outbreak, even with a small number of sexually associated transmissions among those reporting MMSC^15^.

## Conclusion

Using exposure and symptom onset data from 40 monkeypox patients reported in the United States and the Netherlands from May 2022 – June 2022, we estimate the mean incubation period from exposure to first symptom onset of 7.6 days (95% CrI: 6.2–9.7), and using a subset of 21 U.S. monkeypox patients, a mean incubation period from exposure to rash onset of 8.7 days (95% CrI: 6.9–11.7). All 22 U.S. patients included were men, and all reported MMSC, in line with demographic characteristics of people diagnosed with monkeypox in the United Kingdom. As the current outbreak continues to evolve and transmission pathways are clarified, it will be important to determine the incubation period for monkeypox by different routes of exposure, especially if sexual transmission is confirmed. Updated analyses with more data will be important for refining the estimated incubation period of MPXV.

## Data Availability

All data produced in the present work are contained in the manuscript. R code is provided as supplementary material in a Word document.

## Acknowledgements

We thank local, territorial, and state health departments for sharing data about cases as well as laboratories in the Laboratory Reference Network for performing preliminary testing of monkeypox cases for orthopoxvirus. We also thank everyone involved in CDC’s 2022 Monkeypox Response, especially colleagues in CDC’s Division of Global Migration and Quarantine for investigating cases’ travel itineraries. Finally, we thank Miura et al. for making the anonymized data from the Netherlands publicly available.

## Author contributions

Conceptualization: KC

Data curation: KC, NBM, DP, LG, DS, FSM, MAW, CMH, SG, FAR, CB

Formal analysis: KC

Investigation: KC

Methodology: KC

Software: KC

Validation: KC

Visualization: KC

Writing – original draft: KC, NBM

Writing – review & editing: KC, NBM, RBS, LG, DP, SG, FAR, FSM, DS, MAW, YN, CMH, CB

## Footnotes

a No suspicion of other recent Orthopoxvirus exposure (e.g., Vaccinia virus in ACAM2000 vaccination) AND demonstration of the presence of: Orthopoxvirus DNA by polymerase chain reaction of a clinical specimen OR Orthopoxvirus using immunohistochemical or electron microscopy testing methods OR Demonstration of detectable levels of anti-orthopoxvirus IgM antibody during the period of 4 to 56 days after rash onset (https://www.cdc.gov/poxvirus/monkeypox/clinicians/case-definition.html)

b Demonstration of the presence of Monkeypox virus DNA by polymerase chain reaction testing or Next-Generation sequencing of a clinical specimen OR isolation of Monkeypox virus in culture from a clinical specimen (https://www.cdc.gov/poxvirus/monkeypox/clinicians/case-definition.html)

## References

1. World Health Organization. Multi-country monkeypox outbreak: situation update. Disease Outbreak News. Updated 10 June 2022. Accessed 15 June 2022. Available at: https://www.who.int/emergencies/disease-outbreak-news/item/2022-DON392

2. UK Health Security Agency. Monkeypox cases confirmed in England – latest updates. Updated 16 June 2022. Accessed 21 June 2022. Available at: https://www.gov.uk/government/news/monkeypox-cases-confirmed-in-england-latest-updates

3. World Health Organization Regional Office for Europe and European Centre for Disease Prevention and Control. Interim advice for public health authorities on summer events during the monkeypox outbreak in Europe, 2022. Updated 14 June 2022. Accessed 15 June 2022. Available at: https://www.ecdc.europa.eu/sites/default/files/documents/Interim-advice-for-public-health-authorities-on-summer-events-mpx.pdf

4. Nolen LD, Osadebe L, Katomba J, Likofata J, Mukadi D, et al. Extended Human-to-Human Transmission during a Monkeypox Outbreak in the Democratic Republic of the Congo. Emerging Infectious Diseases. 2016,22(6):1014–1021.

5. Breman JG, Kalisa-Ruti Steniowski MV, Zanotto E, Gromyko AI, et al. Human monkeypox, 1970 – 79. Bull World Health Organ. 1980,58(2):165–182.

6. U.S. Centers for Disease Control and Prevention. Signs and Symptoms. Updated June 2022. Accessed 15 June 2022. Available at: https://www.cdc.gov/poxvirus/monkeypox/symptoms.html

7. Reynolds MG, Yorita KL, Kuehnert MJ, Davison WB, Huhn GD, et al. Clinical Manifestations of Human Monkeypox Influenced by Route of Infection. The Journal of Infectious Diseases. 2006;194(6):773–780.

8. Antinori A, Mazzotta V, Vita S, Carletti F, Tacconi D, et al. Epidemiological, clinical and virological characteristics of four cases of monkeypox support transmission through sexual contact, Italy, May 2022. Eurosurveillance. 2022;27(22):pii=2200421.

9. Lessler J, Ott C, Carcelen A, Konikoff J, Williamson J, Bi Q, et al. Times to key events in Zika virus infection and implications for blood donation: a systematic review. Bull World Health Organ. 2016;94:841–9.

10. Miura F, Else van Ewijk C, Backer JA, Xiridou M, Franz E, et al. The incubation period for monkeypox cases confirmed in the Netherlands, May 2022. Euro Surveill. 2022;27(24):pii=2200448.

11. Reich N, Lessler J, Cummings D, Brookmeyer R. Estimating incubation period distributions with coarse data. Stat Med. 2009;28(22):2769–84.

12. Caicedo EY, Charniga K, Rueda A, Dorigatti I, Mendez Y, et al. The epidemiology of Mayaro virus in the Americas: A systematic review and key parameter estimates for outbreak modelling. PLoS Negl Trop Dis. 2021;15(6): e0009418.

13. U.S. Centers for Disease Control and Prevention. Monitoring People Who Have Been Exposed. Updated 8 June 2022. Accessed 15 June 2022. Available at: https://www.cdc.gov/poxvirus/monkeypox/clinicians/monitoring.html

14. UK Health Security Agency. Investigation into monkeypox outbreak in England: technical briefing 1. Updated 10 June 2022. Accessed 15 June 2022. Available at: https://www.gov.uk/government/publications/monkeypox-outbreak-technical-briefings/investigation-into-monkeypox-outbreak-in-england-technical-briefing-1

15. Endo A, Murayama H, Abbott S, Ratnayake R, Pearson CAB, et al. Heavy-tailed sexual contact networks and the epidemiology of monkeypox outbreak in non-endemic regions, May 2022. medRxiv. Updated 13 June 2022. Accessed 15 June 2022. DOI: https://doi.org/10.1101/2022.06.13.22276353

