## Supplemental Code 1 for "Estimating the incubation period of monkeypox virus during the 2022 multi-national outbreak"

**R Code**

\# Estimating incubation periods

\# Updated June 9, 2020 with cases reported up to June 6, 2022

\# N = 22 cases + 18 cases reported from Netherlands = 40

\# Netherlands data:https://www.medrxiv.org/content/10.1101/2022.06.09.22276068v1.full.pdf

\# ASSUMING they report first symptom onset

################################################

\#

\# This analysis uses date of first symptom onset

\#

################################################

\# Creating incubation period dataset

id <- seq(1, 40)

EL <- rep(0, 40)

\# Exposure window (in table)

ER <- c(1, 8, 21, 2, 2, 13, 1, 23, 1, 4, 1, 18, 8, 2, 8, 27, 7, 4, 23, 1, 4, 12, #USA CASES

1, 1, 1, 1, 1, 1, 1, 1, 7, 11, 5, 1, 1, 1, 11, 1, 1, 8) #NETHERLANDS CASES

ER <- ER -0.0007

\# Max days to symptom onset (in table)

SR <- c(6, 8, 23, 5, 6, 14, 11, 24, 12, 6, 8, 18, 16, 2, 17, 29, 10, 6, 23, 3, 8, 12, #USA CASES

7, 10, 9, 9, 7, 7, 15, 20, 7, 16, 20, 8, 6, 7, 17, 13, 9, 10) #NETHERLANDS CASES

SL <- SR - 1

SR <- SR -0.0007

monkeypox <- data.frame(id, EL, ER, SL, SR)

\# Visual summary of interval-censored data

library(dplyr)

library(ggplot2)

library(gridExtra)

dat <- monkeypox %>%

mutate(ELnew = EL-ER,

ERnew = ER-ER,

SLnew = SL-ER,

SRnew = SR-ER)

IP_plot <- ggplot(dat, aes(y=factor(id))) +

geom_segment(aes(x=ELnew, xend=ERnew, yend=factor(id)),

color="blue", size=2) +

geom_segment(aes(x=SLnew, xend=SRnew, yend=factor(id)),

size=2, color="red", alpha=.5) +

ggtitle("Incubation Period data") +

xlab(NULL) +

ylab("id") +

coord_cartesian(xlim = c(-40, 20)) +

theme(axis.text.y = element_text(size=6)) +

annotate("text", x=-35, y="900.1", label="A")

IP_plot

\# The intervals observed for each indivdual.

\# Blue intervals represent windows of possible exposure.

\# Red intervals represent windows of possible time of symptom onset

\# Bayesian MCMC Framework for Estimating Key Distributions

\# the time of infection for each individual, Ei as drawn from a uniform prior

\# defined by the earliest and latest possible times of exposure (ELi and ERi)

\# length of the incubation period, Y is interval censored random variable following a lognormal distribution

\# SLi is the earliest possible time of symptom onset for case i

\# SRi is the latest time of symptom onset for case i

#make a data object for JAGS

monkeypox_jags <- monkeypox

jags_data <- list(

ER=monkeypox_jags$ER,

SL=monkeypox_jags$SL,

SR=monkeypox_jags$SR)

#Let jags know the censoring situation

#1 means interval censored

#2 means event occured after known time

jags_data$IPisCensored=rep(1,40) # no missing

library(rjags)

#define variables to hold the length of the

#time to event for symptoms

jags_data$Y_S <- rep(NA, 40)

#set the initial values for time to event (i.e., Y_S)

IPyInit = jags_data$SL

IPyInit[which(is.na(jags_data$SL)==T)]=0

IPyInit[which(IPyInit==0)]=0.0000000011

#identify which observations are interval censored and which are right censored.

#jags_data$dic=which(jags_data$IPisCensored==1) #this is all incubation period observtions

#set the parameters we want to track

parameters <- c("lm","lsd", "E")

\# specify JAGS model inside R

model1_string <- "

model {

for(i in 1:40){

IPisCensored[i] ~ dinterval( Y_S[i] , IPcensorLimitVec[i,1:2])

IPcensorLimitVec[i,1]<-max(0.000000001,(SL[i]-E[i]))

IPcensorLimitVec[i,2]<-max(0.000000001,(SR[i]-E[i]))

Y_S[i] ~ dlnorm(lm, tau1)

E[i] ~ dunif(0,ER[i])

}

lm~dnorm(0,0.001)

tau1 <-1/lsd^2

lsd~dunif(0,3)

}

"

DistributionFitLWW <-textConnection(model1_string)

set.seed(12345) #if this is not included, multiple initializations maybe needed.

#initialization function for jags

jags_inits <- function() {

rc <-list(E=rep(0.0000000011,40), #start with a fixed E to avoid bad starting points

lm=runif(1,log(2),log(10)),

lsd = runif(1,.1,log(3)),

Y_S=IPyInit)

print(rc)

return(rc)

}

#initialize JAGS model

jagsfit_LWW <- jags.model(DistributionFitLWW,

data=jags_data,

inits=jags_inits,

n.chains=3, quiet=F,

n.adapt=10000)

iters<- 1000000

thin <- 50

full_fit_LWW <- coda.samples(jagsfit_LWW, parameters, n.iter=iters, thin=thin, n.chains=3)

#make all of the chains a single matrix with a burnin removed

ABC1=as.matrix(full_fit_LWW[[1]][,])

ABC2=as.matrix(full_fit_LWW[[2]][,])

ABC3=as.matrix(full_fit_LWW[[3]][,])

ABC1=ABC1[5001:(iters/thin),]

ABC2=ABC2[5001:(iters/thin),]

ABC3=ABC3[5001:(iters/thin),]

#recreate MCMC object for diagnostics

full_fit_LWW<-list(as.mcmc(ABC1), as.mcmc(ABC2), as.mcmc(ABC3))

chains_LWW <- rbind(ABC1,ABC2,ABC3)

colnames(chains_LWW) <- varnames(full_fit_LWW[[1]])

chains_LWW <- as.data.frame(chains_LWW)

#save(full_fit_LWW, chains_LWW, file="../full_fit_jags_LWW.RData")

\# $\hat{R}$ statistic for primary results

#load("../full_fit_jags_LWW.RData")

print(gelman.diag(full_fit_LWW))

\# Estimates of key distributions

\# Distribution of parameters and key quantiles of the incubation period for MPXV infection.

require(knitr)

inc_fit_jags_LWW <- quantile(exp(chains_LWW$lm+chains_LWW$lsd^2/2), prob=c(0.5,0.025,0.975))

inc_fit_jags_LWW <- rbind(inc_fit_jags_LWW,

exp(c(median(chains_LWW$lm), quantile(chains_LWW$lm,prob=c(0.025,0.975)))))

inc_fit_jags_LWW <- rbind(inc_fit_jags_LWW,

exp(c(mean(chains_LWW$lsd), quantile(chains_LWW$lsd,prob=c(0.025,0.975)))))

inc_fit_jags_LWW <- rbind(inc_fit_jags_LWW,

quantile(sqrt((exp(chains_LWW$lsd^2)-1)*exp(2*chains_LWW$lm + chains_LWW$lsd^2)),

prob=c(0.5,0.025,0.975)))

for (q in c(0.05, 0.25, 0.5, 0.75, 0.95)) {

tmp <- qlnorm(q, chains_LWW$lm, chains_LWW$lsd)

inc_fit_jags_LWW <- rbind(inc_fit_jags_LWW,

c(mean(tmp), quantile(tmp, prob=c(0.025, 0.975))))

}

colnames(inc_fit_jags_LWW) <- c("est","CIlow","CIhigh")

rownames(inc_fit_jags_LWW) <- c("mean","median",

"dispersion",

"sd",

"p5","p25","p50","p75","p95")

kable(inc_fit_jags_LWW, format="markdown", digits=2)

\# Plot the estimates

#first make data frame to hold everything

inc_curve <- NULL

for (q in seq(0,60,.1)) {

tmp <- plnorm(q, chains_LWW$lm, chains_LWW$lsd)

tmp <- quantile(tmp, prob=c(0.025, .5, 0.975))

inc_curve <- rbind(inc_curve, c(q=q,

plow=tmp[1],

pmid=tmp[2],

phigh=tmp[3]))

}

inc_curve <- as.data.frame(inc_curve)

colnames(inc_curve) <- c("q","incplow","incpmid","incphigh")

#require(ggplot2)

inc_plt <- ggplot(inc_curve, aes(x=q)) +

geom_ribbon(aes(ymin=incplow, ymax=incphigh), fill="blue", alpha=.2) +

geom_line(aes(y=incpmid), col="blue") +theme_bw()+

scale_x_continuous(limits=c(0, 60), expand = c(0, 0)) +

ylab("Proportion of cases who had symptoms") + xlab("Time from exposure to MPXV (days)")

inc_plt

\# Estimating incubation periods

\# Updated June 10, 2020 with cases reported up to June 6, 2022

\# N = 21 cases

################################################

\#

\# This analysis uses date of rash onset

\#

################################################

\# Creating incubation period dataset

id <- seq(1, 21)

EL <- rep(0, 21)

ER <- c(1, 10, 21, 2, 2, 13, 1, 23, 1, 4, 1, 18, 8, 7, 8, 27, 7, 4, 1, 4, 14) # Exposure window (in table)

ER <- ER -0.0007

SR <- c(6, 11, 28, 5, 6, 15, 11, 24, 12, 12, 8, 18, 16, 9, 18, 36, 10, 7, 13, 18, 17) # Max days to symptom onset (in table)

SL <- SR - 1

SR <- SR -0.0007

monkeypox <- data.frame(id, EL, ER, SL, SR)

\# Visual summary of interval-censored data

library(dplyr)

library(ggplot2)

library(gridExtra)

dat <- monkeypox %>%

mutate(ELnew = EL-ER,

ERnew = ER-ER,

SLnew = SL-ER,

SRnew = SR-ER)

IP_plot <- ggplot(dat, aes(y=factor(id))) +

geom_segment(aes(x=ELnew, xend=ERnew, yend=factor(id)),

color="blue", size=2) +

geom_segment(aes(x=SLnew, xend=SRnew, yend=factor(id)),

size=2, color="red", alpha=.5) +

ggtitle("Incubation Period data") +

xlab(NULL) +

ylab("id") +

coord_cartesian(xlim = c(-40, 20)) +

theme(axis.text.y = element_text(size=6)) +

annotate("text", x=-35, y="900.1", label="A")

IP_plot

\# The intervals observed for each indivdual.

\# Blue intervals represent windows of possible exposure.

\# Red intervals represent windows of possible time of symptom onset

\# Bayesian MCMC Framework for Estimating Key Distributions

\# the time of infection for each individual, Ei as drawn from a uniform prior

\# defined by the earliest and latest possible times of exposure (ELi and ERi)

\# length of the incubation period, Y is interval censored random variable following a lognormal distribution

\# SLi is the earliest possible time of symptom onset for case i

\# SRi is the latest time of symptom onset for case i

#make a data object for JAGS

monkeypox_jags <- monkeypox

jags_data <- list(

ER=monkeypox_jags$ER,

SL=monkeypox_jags$SL,

SR=monkeypox_jags$SR)

#Let jags know the censoring situation

#1 means interval censored

#2 means event occured after known time

jags_data$IPisCensored=rep(1,21) # no missing

library(rjags)

#define variables to hold the length of the

#time to event for symptoms

jags_data$Y_S <- rep(NA, 21)

#set the initial values for time to event (i.e., Y_S)

IPyInit = jags_data$SL

IPyInit[which(is.na(jags_data$SL)==T)]=0

IPyInit[which(IPyInit==0)]=0.0000000011

#identify which observations are interval censored and which are right censored.

#jags_data$dic=which(jags_data$IPisCensored==1) #this is all incubation period observtions

#set the parameters we want to track

parameters <- c("lm","lsd", "E")

\# specify JAGS model inside R

model1_string <- "

model {

for(i in 1:21){

IPisCensored[i] ~ dinterval( Y_S[i] , IPcensorLimitVec[i,1:2])

IPcensorLimitVec[i,1]<-max(0.000000001,(SL[i]-E[i]))

IPcensorLimitVec[i,2]<-max(0.000000001,(SR[i]-E[i]))

Y_S[i] ~ dlnorm(lm, tau1)

E[i] ~ dunif(0,ER[i])

}

lm~dnorm(0,0.001)

tau1 <-1/lsd^2

lsd~dunif(0,3)

}

"

DistributionFitLWW <-textConnection(model1_string)

set.seed(12345) #if this is not included, multiple initializations maybe needed.

#initialization function for jags

jags_inits <- function() {

rc <-list(E=rep(0.0000000011,21), #start with a fixed E to avoid bad starting points

lm=runif(1,log(2),log(10)),

lsd = runif(1,.1,log(3)),

Y_S=IPyInit)

print(rc)

return(rc)

}

#initialize JAGS model

jagsfit_LWW <- jags.model(DistributionFitLWW,

data=jags_data,

inits=jags_inits,

n.chains=3, quiet=F,

n.adapt=10000)

iters<- 1000000

thin <- 50

full_fit_LWW <- coda.samples(jagsfit_LWW, parameters, n.iter=iters, thin=thin, n.chains=3)

#make all of the chains a single matrix with a burnin removed

ABC1=as.matrix(full_fit_LWW[[1]][,])

ABC2=as.matrix(full_fit_LWW[[2]][,])

ABC3=as.matrix(full_fit_LWW[[3]][,])

ABC1=ABC1[5001:(iters/thin),]

ABC2=ABC2[5001:(iters/thin),]

ABC3=ABC3[5001:(iters/thin),]

#recreate MCMC object for diagnostics

full_fit_LWW<-list(as.mcmc(ABC1), as.mcmc(ABC2), as.mcmc(ABC3))

chains_LWW <- rbind(ABC1,ABC2,ABC3)

colnames(chains_LWW) <- varnames(full_fit_LWW[[1]])

chains_LWW <- as.data.frame(chains_LWW)

#save(full_fit_LWW, chains_LWW, file="../full_fit_jags_LWW.RData")

\# $\hat{R}$ statistic for primary results

#load("../full_fit_jags_LWW.RData")

print(gelman.diag(full_fit_LWW))

\# Estimates of key distributions

\# Distribution of parameters and key quantiles of the incubation period for MPXV infection.

require(knitr)

inc_fit_jags_LWW <- quantile(exp(chains_LWW$lm+chains_LWW$lsd^2/2), prob=c(0.5,0.025,0.975))

inc_fit_jags_LWW <- rbind(inc_fit_jags_LWW,

exp(c(median(chains_LWW$lm), quantile(chains_LWW$lm,prob=c(0.025,0.975)))))

inc_fit_jags_LWW <- rbind(inc_fit_jags_LWW,

exp(c(mean(chains_LWW$lsd), quantile(chains_LWW$lsd,prob=c(0.025,0.975)))))

inc_fit_jags_LWW <- rbind(inc_fit_jags_LWW,

quantile(sqrt((exp(chains_LWW$lsd^2)-1)*exp(2*chains_LWW$lm + chains_LWW$lsd^2)),

prob=c(0.5,0.025,0.975)))

for (q in c(0.05, 0.25, 0.5, 0.75, 0.95)) {

tmp <- qlnorm(q, chains_LWW$lm, chains_LWW$lsd)

inc_fit_jags_LWW <- rbind(inc_fit_jags_LWW,

c(mean(tmp), quantile(tmp, prob=c(0.025, 0.975))))

}

colnames(inc_fit_jags_LWW) <- c("est","CIlow","CIhigh")

rownames(inc_fit_jags_LWW) <- c("mean","median",

"dispersion",

"sd",

"p5","p25","p50","p75","p95")

kable(inc_fit_jags_LWW, format="markdown", digits=2)

\# Plot the estimates

#first make data frame to hold everything

inc_curve <- NULL

for (q in seq(0,60,.1)) {

tmp <- plnorm(q, chains_LWW$lm, chains_LWW$lsd)

tmp <- quantile(tmp, prob=c(0.025, .5, 0.975))

inc_curve <- rbind(inc_curve, c(q=q,

plow=tmp[1],

pmid=tmp[2],

phigh=tmp[3]))

}

inc_curve <- as.data.frame(inc_curve)

colnames(inc_curve) <- c("q","incplow","incpmid","incphigh")

#require(ggplot2)

inc_plt <- ggplot(inc_curve, aes(x=q)) +

geom_ribbon(aes(ymin=incplow, ymax=incphigh), fill="blue", alpha=.2) +

geom_line(aes(y=incpmid), col="blue") +theme_bw()+

scale_x_continuous(limits=c(0, 60), expand = c(0, 0)) +

ylab("Proportion of cases who had symptoms") + xlab("Time from exposure to MPXV (days)")

inc_plt
